# Knowledge, Attitudes, and Practices (KAP) Regarding Photoprotection Among Medical Students at a Nicaraguan University: A Cross-Sectional Study

**DOI:** 10.64898/2026.02.24.26347045

**Authors:** Jerha Montes, Britthany Noguera, Ariana Obregón, Alejandro Rivas, Hallet Whynot, Ruby Poveda, Virgilio Blandón

**Affiliations:** Medical Shool, Universidad Iberoamericana de Ciencias y Tecnología (UNICIT), Managua, Nicaragua; Department of Dermatology, Hospital Carlos Roberto Huembes, Managua, Nicaragua; Faculty of Medicine, Universidad Iberoamericana de Ciencias y Tecnología (UNICIT), Managua, Nicaragua

**Keywords:** Photoprotection, Sunscreening agents, Health knowledge, attitudes, practice, Medical students, Skin cancer, Nicaragua

## Abstract

**Background:** Medical students represent a critical population for photoprotection education, as future physicians responsible for skin cancer prevention counseling. However, no previous studies have characterized knowledge, attitudes, and practices (KAP) regarding photoprotection among medical students in Central America or the Caribbean.

**Objective:** To assess KAP related to photoprotection and identify associated factors among medical students at a Nicaraguan university.

**Methods:** A cross-sectional study was conducted among 133 medical students at the Universidad Iberoamericana de Ciencias y Tecnología (UNICIT), Managua, Nicaragua. An ad hoc questionnaire assessed sociodemographic characteristics, knowledge, attitudes, and photoprotective practices. Domain-specific and global KAP scores were calculated. Bivariate analyses examined associations with sex, academic year, skin phototype, and age.

**Results:** Participants were predominantly female (73.7%), with a median age of 20 years (IQR: 18–21). Although 97.0% knew what sunscreen is and 88.0% correctly identified adequate sunscreen characteristics, only 33.1% knew the minimum recommended SPF for daily use, and 21.8% understood endogenous photoprotective mechanisms. Regular sunscreen use was reported by 39.1%, while 24.8% reported never using it. Women demonstrated significantly higher scores across all domains, with moderate effect sizes for practice (d = 0.56) and global KAP (d = 0.60). No improvements were observed across academic years (p > 0.05). Age showed weak negative correlations with practice (ρ = −0.237; p = 0.006) and global KAP (ρ = −0.204; p = 0.018). The primary barrier to sunscreen use was forgetfulness (49.6%).

**Conclusions:** This first KAP study among medical students in Nicaragua reveals a substantial gap between photoprotection knowledge and practice. Current medical training appears insufficient to promote sustained protective behaviors. Findings support integrating practical, behavior-oriented photoprotection education into medical curricula and establish a regional baseline for future interventions.

## 1. Introduction

Exposure to solar ultraviolet radiation (UVR) is a well-recognized environmental carcinogen and a major modifiable risk factor for cutaneous malignancies, including melanoma and non-melanoma skin cancers [1,2], as well as for premature skin aging [3]. UVR induces DNA damage through both direct absorption (primarily UVB) and oxidative stress (primarily UVA), and its role in skin carcinogenesis has been confirmed by epidemiological, clinical, and molecular studies, with factors such as phototype and melanin content influencing individual susceptibility [4]. Photoprotection comprises a spectrum of preventive behaviors, such as regular application of sunscreen, avoidance of peak sun exposure, seeking shade, and the use of protective clothing, which are fundamental to reduce the harmful effects of UVR [5].

Medical students constitute a critical population for evaluating knowledge, attitudes, and practices (KAP) related to photoprotection. As future healthcare providers, they are expected not only to adopt effective sun-protective behaviors themselves but also to actively educate patients and the broader community about strategies for preventing skin cancer [6]. Previous studies conducted in countries such as Peru, Vietnam, Italy, and Spain have consistently shown that medical students often possess adequate theoretical knowledge about photoprotection; however, this knowledge frequently does not translate into consistent preventive practices [7–9]. These international studies are cited to provide comparative evidence and to highlight that gaps between knowledge and practice among medical students are not unique to one region but may represent a widespread challenge in medical education.

In Latin America, countries located mainly in intertropical zones experience high year-round solar irradiance. The region also exhibits significant ethnic diversity, resulting in a wide range of skin phototypes [10]. Socioeconomic factors, including education, income, and access to health services, also influence photoprotection behaviors, as populations with fewer resources or limited knowledge about sun protection are less likely to adopt preventive measures against UV-related skin damage [11,12]. These factors are important to consider in studies of photoprotection.

To our knowledge, no published research has assessed photoprotection KAP among medical students in Central America and Caribbean. In response to this gap, this exploratory study was conducted to assess photoprotection KAP among medical students at a Nicaraguan university using an ad hoc questionnaire. The primary objective was to identify specific areas of insufficient knowledge, prevailing behavioral patterns, and perceived barriers to effective photoprotection. The results are intended to inform the development of targeted educational strategies within medical curricula and to support future national efforts in skin cancer prevention.

## 2. Methods

### Study Design

This was a quantitative, cross-sectional, exploratory study designed to characterize the sociodemographic profile of medical students at the Universidad Iberoamericana de Ciencia y Tecnología (UNICIT) and to assess their knowledge, attitudes, and practices (KAP) regarding photoprotection and skin cancer prevention. The study also aimed to identify behavioral patterns, explore associated factors, and describe perceived barriers related to photoprotective practices. Given the limited prior research on this topic within the local student population, an ad hoc instrument was developed to capture context-specific dimensions relevant to this setting.

### Study Population and Sample

The study population included all medical students enrolled in the Faculty of Medical Sciences during the academic period of data collection (N = 210). Sample size was calculated assuming a 95% confidence level, a 5% margin of error, and an expected proportion of 50%, resulting in a target sample of 136 participants. A total of 133 students completed the survey, corresponding to a confidence level of 94.26% and a margin of error of 5.17%. Participation was voluntary.

### Variables and Operational Definitions

Sociodemographic variables included age, gender, place of origin, academic year, sefl-perceived skin type and skin phototype classified according to the Fitzpatrick scale (I–VI) (Figure S1).

The primary outcome variables were knowledge, attitudes, and practices related to photoprotection. These domains were assessed through 19 structured items. The knowledge domain included items on conceptual understanding of photoprotection, recommended SPF levels, characteristics of adequate sunscreen, environmental impact, and endogenous photoprotective mechanisms. The attitudes domain explored perceptions of risk, motivation for sunscreen use, verification behaviors (e.g., reading ingredients and checking SPF), and criteria for product selection. The practices domain assessed frequency of sunscreen use, SPF level, reapplication habits, use during outdoor activities, age of initiation, family practices, use of additional protective measures, and recommendation behaviors.

Each item contributed to a predefined scoring system. Responses were coded and standardized to a 0–1 scale to ensure comparability across items. Domain scores were calculated as follows: Knowledge (range 0–6), Attitudes (range 0–6), and Practices (range 0–7). The global KAP score ranged from 0 to 19 and represented the sum of the three domains. Higher scores indicated better knowledge, more favorable attitudes, and more consistent photoprotective practices. Aggregated scores were primarily used for descriptive and bivariate analyses and were not intended for predictive modeling. Variables related to barriers and facilitating factors were analyzed descriptively and were not included in composite scores.

### Data Collection Instrument

Data were collected using a self-administered, structured survey developed specifically for this study based on an extensive literature review on KAP studies in photoprotection and skin cancer prevention. Content validity was assessed through expert review by a board-certified dermatologist and a dermatology resident, who evaluated clarity, relevance, and conceptual alignment of items with study objectives.

A pilot test was conducted with 15 medical students to assess comprehensibility and response flow. Minor wording adjustments were made accordingly. Participants involved in the pilot phase were excluded from the final analysis.

As this was an ad hoc instrument developed for exploratory purposes, formal psychometric validation, including construct validity and reliability testing, was not performed. Therefore, comparisons with standardized KAP instruments should be interpreted with caution. The complete survey instrument, including item structure, scoring criteria, and coding procedures, is provided in the Supplementary Data (Table S1-2).

### Data Collection Procedure

The survey was distributed in person during academic activities after authorization from institutional authorities. Participants received a brief explanation of the study objectives and provided informed consent prior to completion. The questionnaire was anonymous and required approximately 10–15 minutes to complete.

### Ethical Considerations

This study was conducted in accordance with the ethical principles of the Declaration of Helsinki. The research protocol was reviewed and approved by the academic authorities of Universidad Iberoamericana de Ciencias y Tecnología (UNICIT). All participants provided written informed consent prior to participation. The survey was anonymous, self-administered, and confidential; no personally identifiable information was collected. The consent form is available in the Supplementary Materials. Data are reported exclusively in aggregate form to ensure participant anonymity.

### Data Management and Missing Data

Data were entered and analyzed using IBM SPSS Statistics version 27. Data cleaning procedures included verification of coding accuracy and range checks. Missing data were minimal, with less than 5% missing for most variables. The variable “age of sunscreen initiation” had 16 missing values (12%), corresponding to participants who reported never having used sunscreen; for analytical purposes, this was handled using a categorized alternative variable distinguishing users from non-users (Table S3).

Outliers were assessed using the interquartile range method. Identified extreme values were retained, as they were consistent with plausible variability within the study population and did not reflect data entry errors.

### Statistical Analysis

Descriptive statistics were calculated for all variables. Categorical data are presented as frequencies and percentages. Continuous variables were summarized using means and standard deviations when normally distributed, or medians and interquartile ranges otherwise.

Distributional assumptions were evaluated using the Kolmogorov–Smirnov test with Lilliefors correction and inspection of Q–Q plots (Table S4, Figure S2–4). Homogeneity of variances was assessed with Levene’s test (Table S5). When normality was met but variance homogeneity was violated, the Brown–Forsythe robust ANOVA was applied. Independence of observations was ensured by the anonymous, self-administered design.

Given the exploratory nature of the study and the use of an ad hoc instrument, analyses were restricted to bivariate comparisons. Depending on distributional assumptions, group differences were examined using Student’s t-test or one-way ANOVA for parametric data, and Mann–Whitney U or Kruskal–Wallis tests for non-parametric data. Post hoc analyses were performed only when overall tests were significant, using Bonferroni or Games–Howell corrections according to variance assumptions. Associations between age and KAP scores were assessed using Spearman’s rank correlation.

Because the instrument was not formally validated, individual photoprotection behaviors contributing to total Practice and total KAP scores were additionally examined descriptively to evaluate internal coherence of the scoring structure. To avoid analytic circularity, these exploratory analyses are reported exclusively in the Supplementary Materials (Table S6, Figure S5).

Multivariable regression modeling was not performed, as the primary objective was descriptive rather than predictive. Statistical significance was set at p < 0.05; however, interpretation prioritized effect size, directionality, and consistency of patterns in light of the cross-sectional design and measurement limitations.

### Additional Methodology: Exploratory Literature Search

To contextualize the novelty of this study, an exploratory literature search was conducted after data collection to identify previous studies on knowledge, attitudes, and practices (KAP) regarding photoprotection in medical students from Central America and the Caribbean. Searches across PubMed, SciELO, Dialnet, DOAJ, and Google Scholar found 18 relevant studies from other regions (South America, Europe, Asia, North America, Africa), but none from Central America or the Caribbean, confirming a regional evidence gap (Figure S6, Table S6-7)

## 3. Results

### 3.1. Sociodemographic Characteristics

A total of 133 medical students were included in the analysis. The median age was 20 years (IQR: 18–21; range: 16–28), reflecting a predominantly young population (Supplementary Figure S1). Women represented 73.7% of the sample (n = 98), while men accounted for 26.3% (n = 35). Most participants were from Managua (67.7%), followed by Masaya (8.3%) and Granada (6.0%). The largest proportion of students were enrolled in the fourth academic year (31.6%), followed by first (22.6%) and third year (21.8%).

Regarding skin phototype, types III (36.4%), II (25.0%), and IV (22.0%) were the most frequent. In terms of self-perceived skin type, combination skin was the most commonly reported type (36.1%; n = 48), followed by oily skin (31.6%; n = 42). A smaller proportion of the sample reported sensitive (9.0%; n = 12) or dry skin (7.5%; n = 10), while 15.8% (n = 21) were unaware of their skin type. Detailed characteristics are presented in Table 1.

**Table 1.**
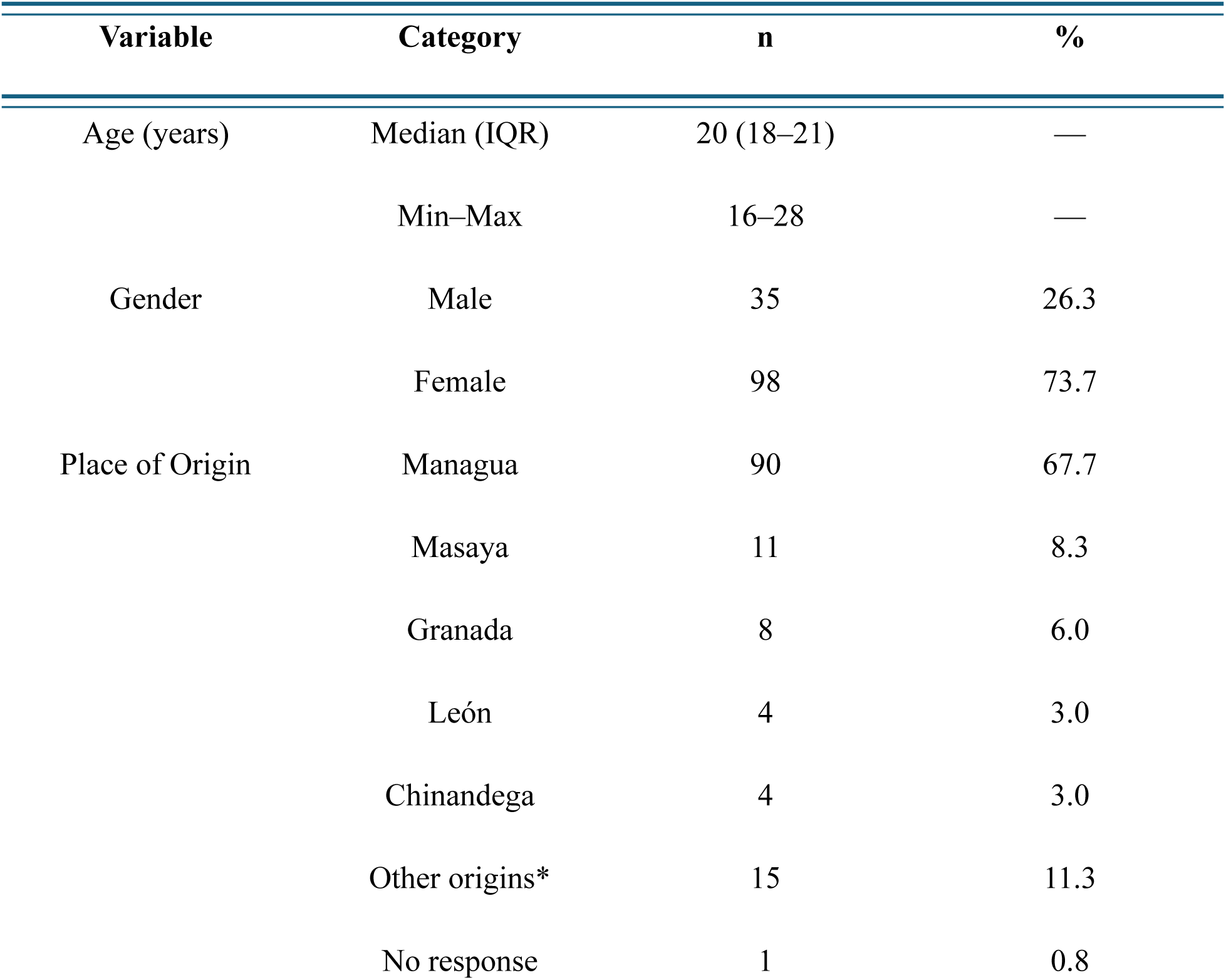
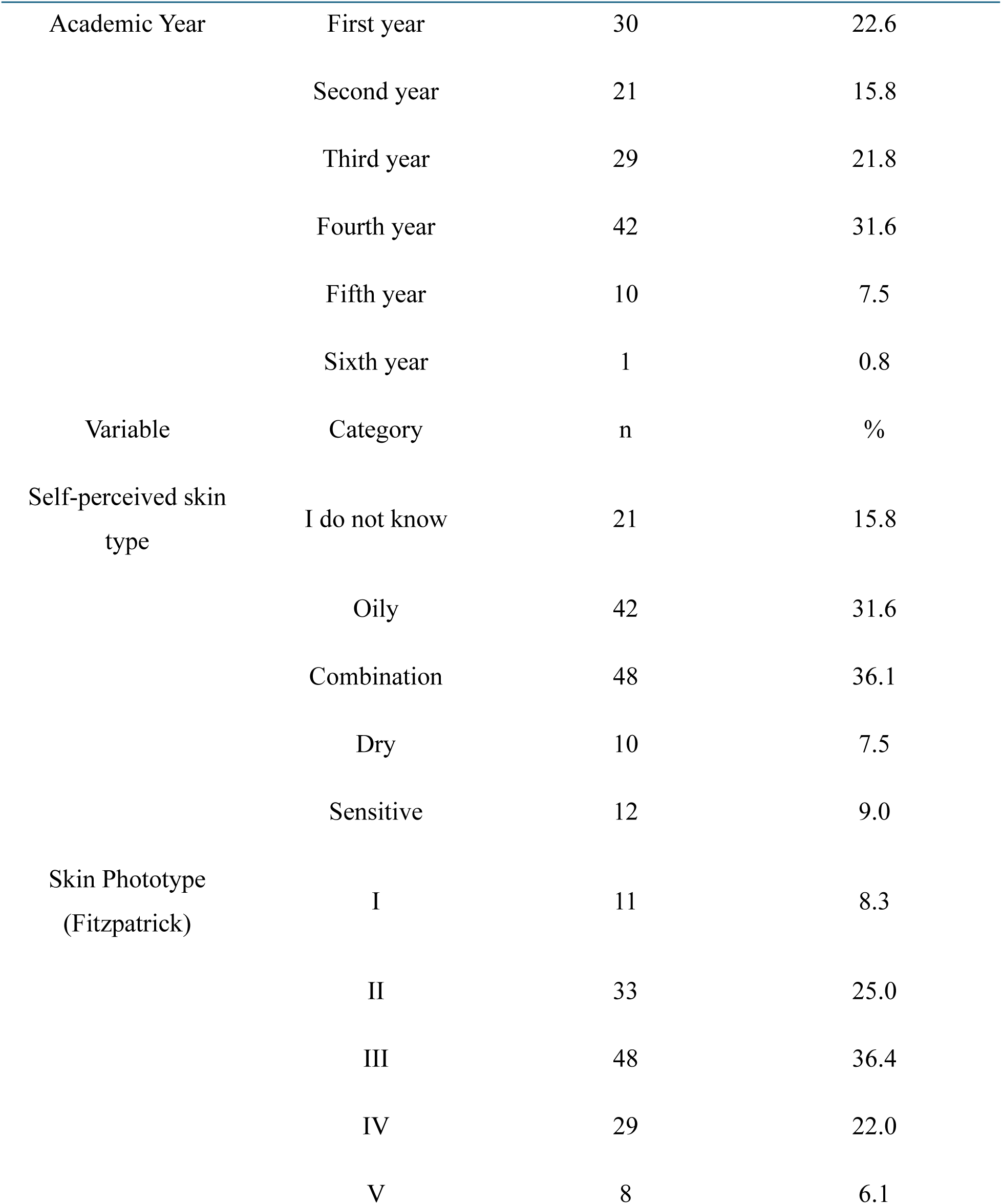
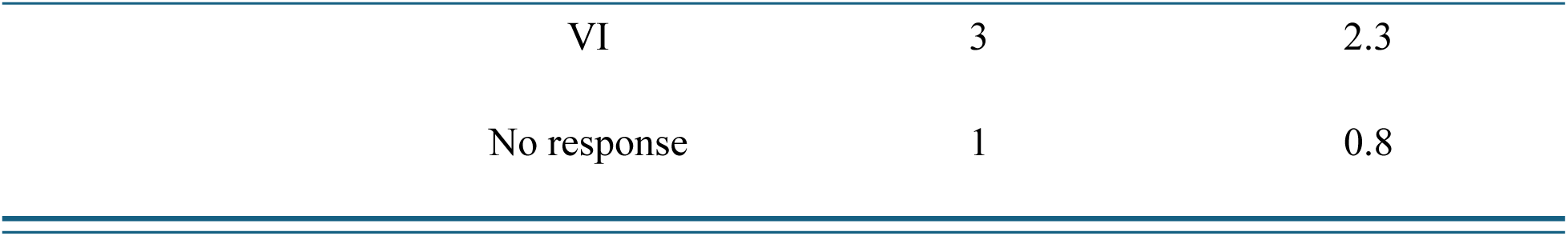
Sociodemographic and Clinical Characteristics of Participants (n = 133) Data are presented as absolute frequency (n) and percentage (%), except for age, which is reported as median and interquartile range (IQR) due to non-normal distribution. Skin phototype was classified according to the Fitzpatrick scale. IQR: interquartile range.

### 3.2. Knowledge, Attitudes, and Practices Toward Photoprotection

In the knowledge domain, most participants reported knowing what sunscreen is (97.0%); however, only 60.9% indicated understanding the concept of photoprotection. Knowledge regarding the minimum recommended SPF for daily use was limited, with 33.1% answering correctly. While 88.0% correctly identified the characteristics of an adequate sunscreen, awareness of environmental impact (30.1%) and endogenous photoprotective mechanisms such as melanin (21.8%) was notably lower.

Regarding practices, regular sunscreen use was reported by 39.1% of participants, whereas 36.1% used it occasionally and 24.8% reported never using it. Most students (86.5%) indicated using SPF ≥30. The median age of sunscreen initiation was 16 years (IQR: 12.5–18), and 12.0% reported never having used sunscreen. Nearly half of the participants (48.9%) reported regular sunscreen use within their family environment. Additionally, 55.6% reported using at least one additional photoprotective measure, and 89.5% indicated that they consistently recommend sunscreen use to others.

In terms of attitudes, only 34.6% of students reported having sufficient information about the risks associated with excessive sun exposure. Verification behaviors were inconsistent: 27.8% always read sunscreen ingredients and 27.1% always checked SPF prior to use. The primary motivation for sunscreen use was combined prevention (40.6%), followed by prevention of sunburn (27.1%). When selecting a sunscreen, 63.9% reported considering multiple factors, and cream was the preferred formulation (63.9%).

Complete item-level distributions for knowledge, attitudes, and practices are provided in Table 2.

**Table 2.**
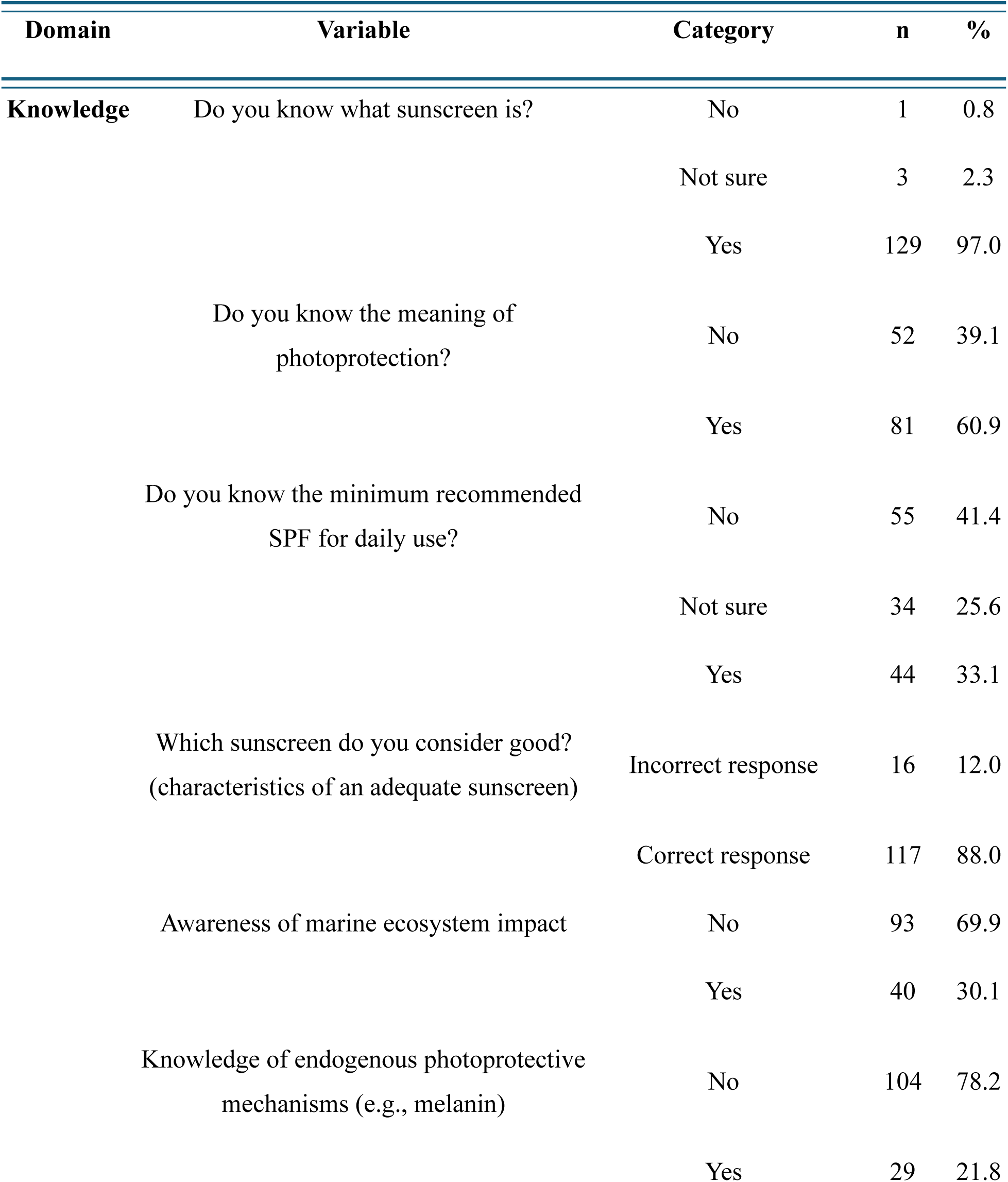
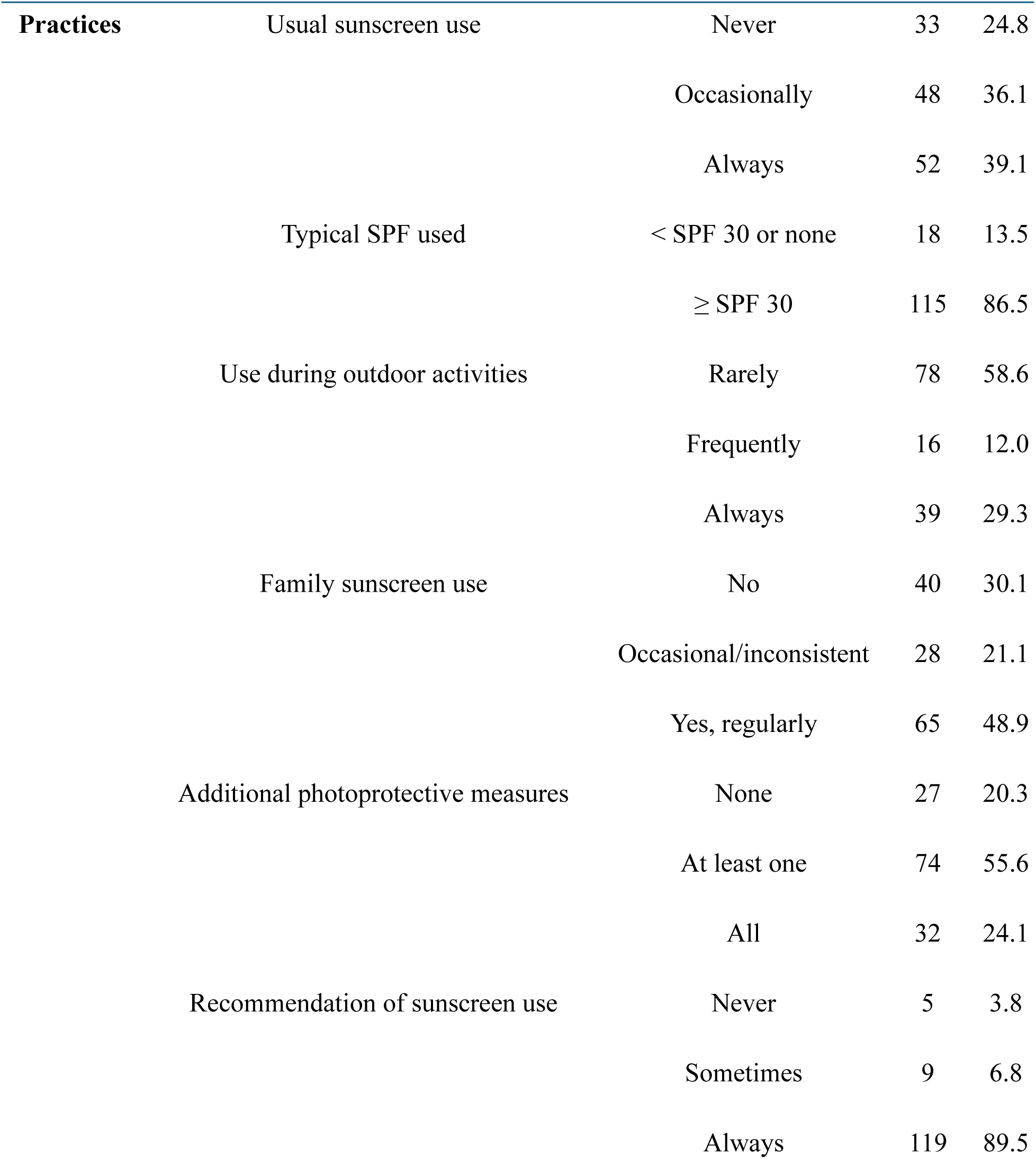
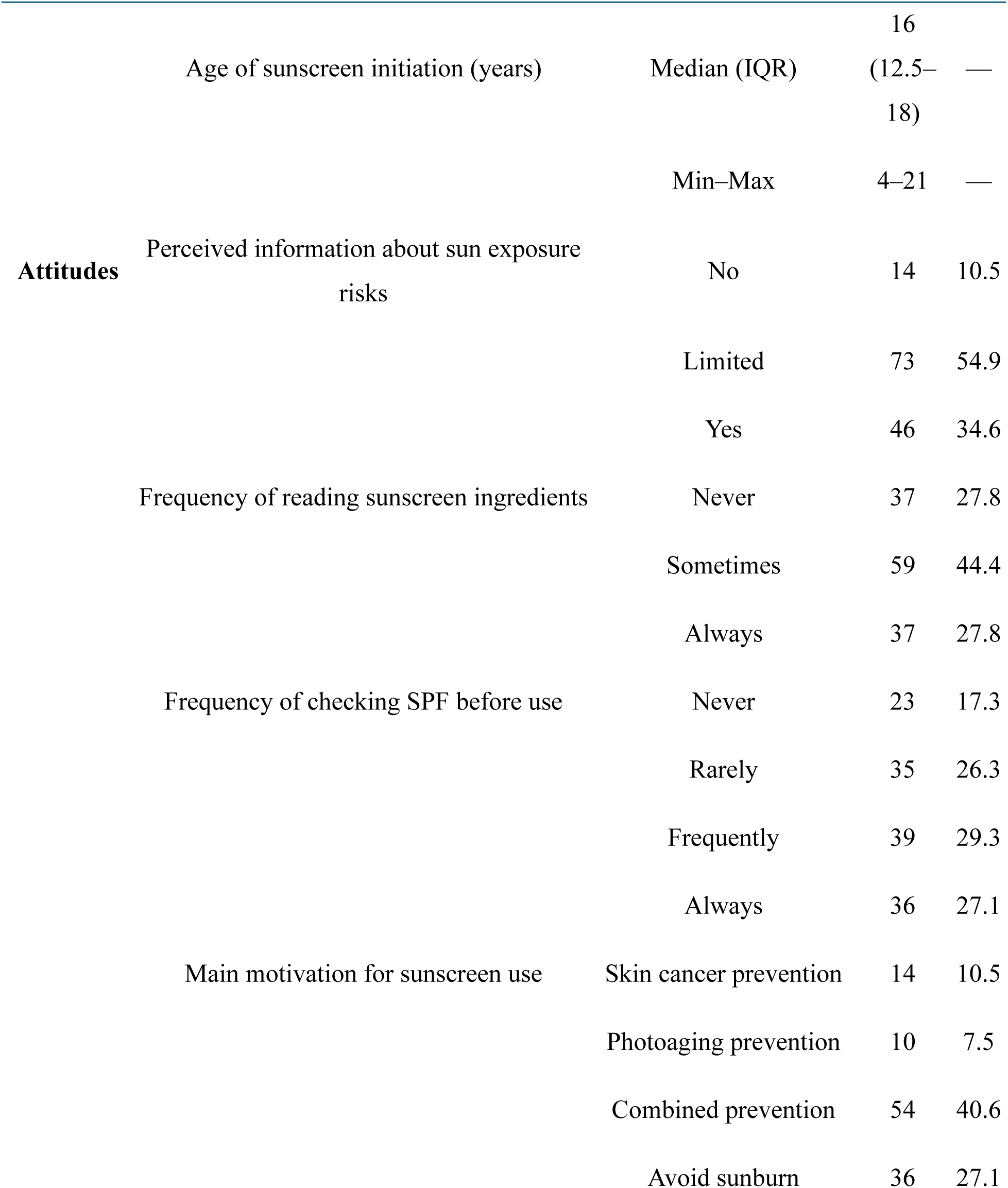
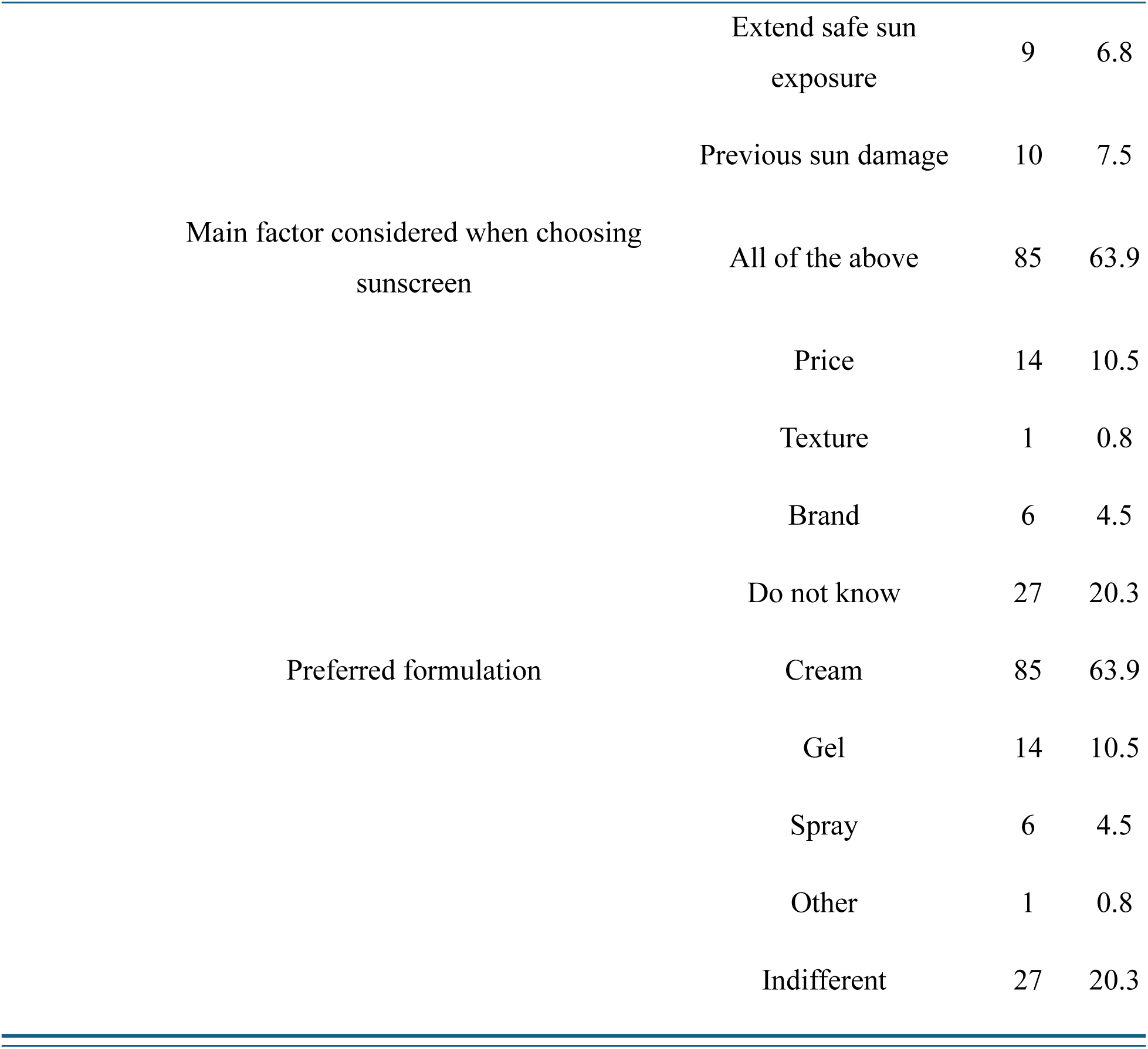
Distribution of Knowledge, Attitudes, and Practices Toward Photoprotection Among Participants (n = 133) Data are presented as absolute frequencies (n) and percentages (%). Percentages were calculated based on valid responses. Age of sunscreen initiation is reported as median and interquartile range (IQR) with minimum and maximum values due to non-normal distribution

### 3.3. Global KAP Scores and Distributional Assumptions

Normality was assessed using the Kolmogorov–Smirnov test with Lilliefors correction and visual inspection of Q–Q plots for continuous variables, including age, age of sunscreen initiation, and KAP domain scores.

Age of participants and age of sunscreen initiation did not follow a normal distribution and are therefore reported as medians with interquartile ranges. Similarly, Knowledge and Attitude scores were non-normally distributed and are presented as medians (Knowledge: 3.5, IQR: 2.5–4.0; Attitudes: 4.17, IQR: 3.33–4.67).

In contrast, Practice scores (mean 5.04 ± 1.24) and global KAP scores (mean 12.4271 ± 2.52) approximated normal distributions and are reported as means with standard deviations.

Homogeneity of variances was evaluated using Levene’s test for group comparisons. Assumptions were met in most analyses; when violations occurred, the Brown–Forsythe correction was applied. Detailed results of normality and homogeneity testing are presented in Supplementary data (Table S4, Figure S2-3)

### 3.4. Sociodemographics and KAP Associations

Women exhibited significantly higher Knowledge, Attitude, Practice, and global KAP scores compared to men (all p < 0.05), with moderate effect sizes observed for Practice (d = 0.56) and overall KAP scores (d = 0.60). No significant differences were found in any KAP domain according to academic year or skin phototype (all p > 0.05) (Figure 1).

**Figure 1.**
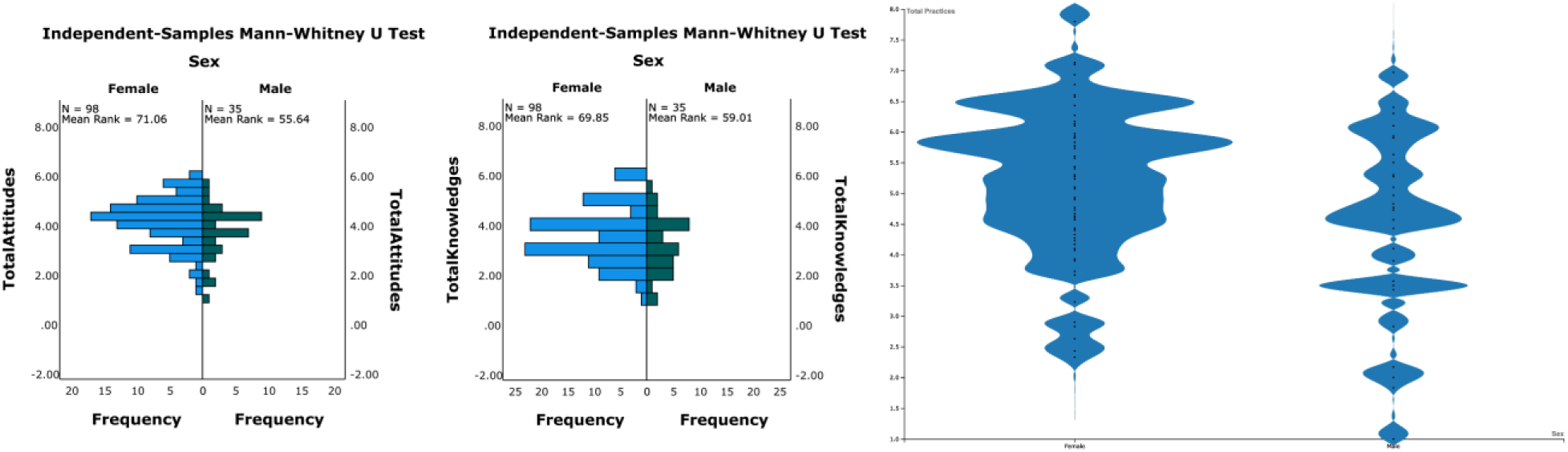
Comparison of Knowledge, Attitude, and Practice (KAP) scores by sex. Left and center panels display independent-samples Mann–Whitney U tests comparing total Knowledge and total Attitude scores between males (n=35) and females (n=98), showing significant differences in mean ranks (all p < 0.05). The right panel illustrates the distribution of total Practice scores between sexes; women exhibited significantly higher Practice scores (p = 0.005) with a moderate effect size (Cohen’s d = 0.56).

Age showed weak but statistically significant negative correlations with Practice (ρ = −0.237; p = 0.006) and global KAP score (ρ = −0.204; p = 0.018), whereas correlations with Knowledge and Attitude were not significant.

All associations between sociodemographic variables, specific practices, and KAP scores are summarized in Table 3.

**Table 3.**
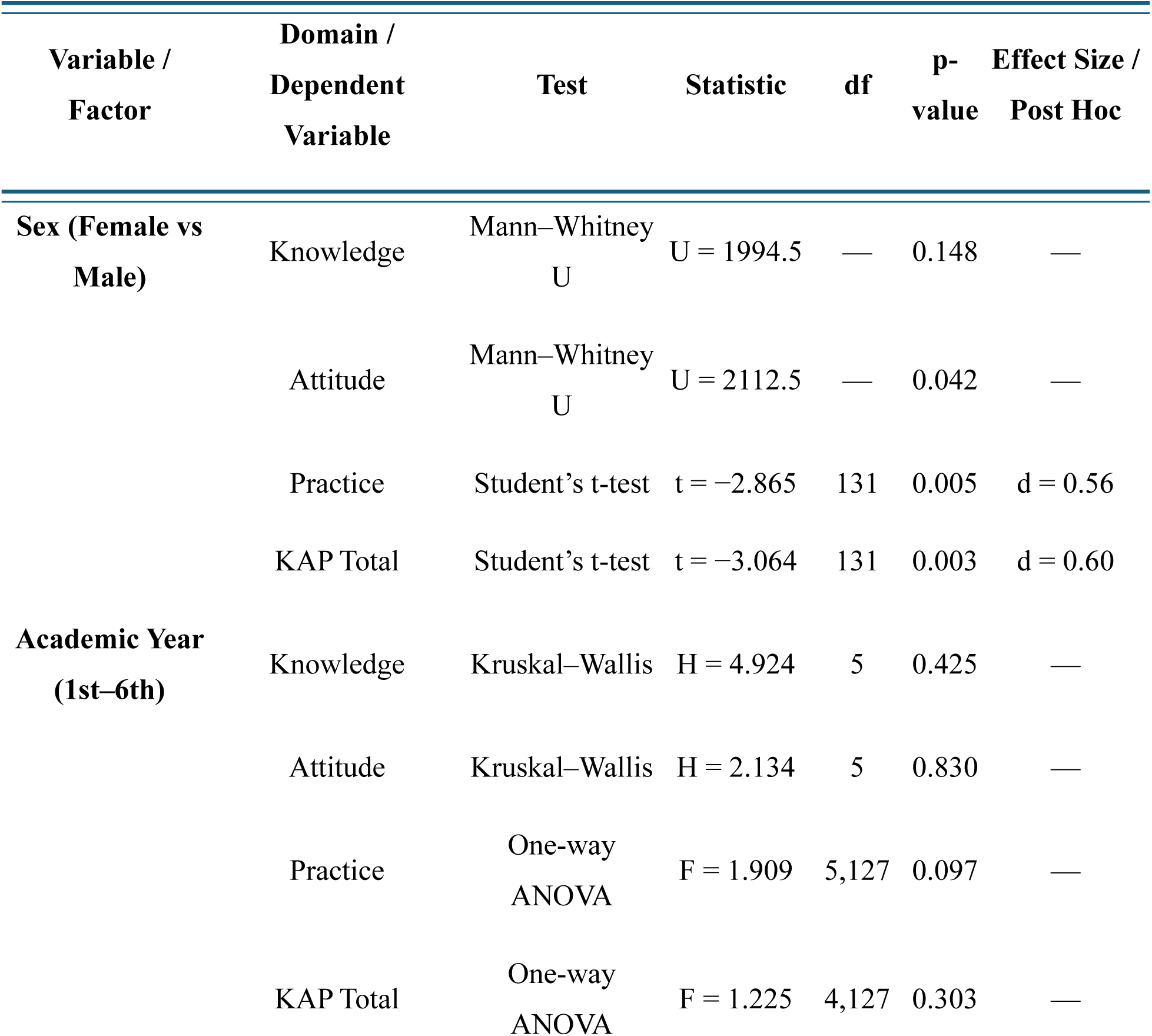
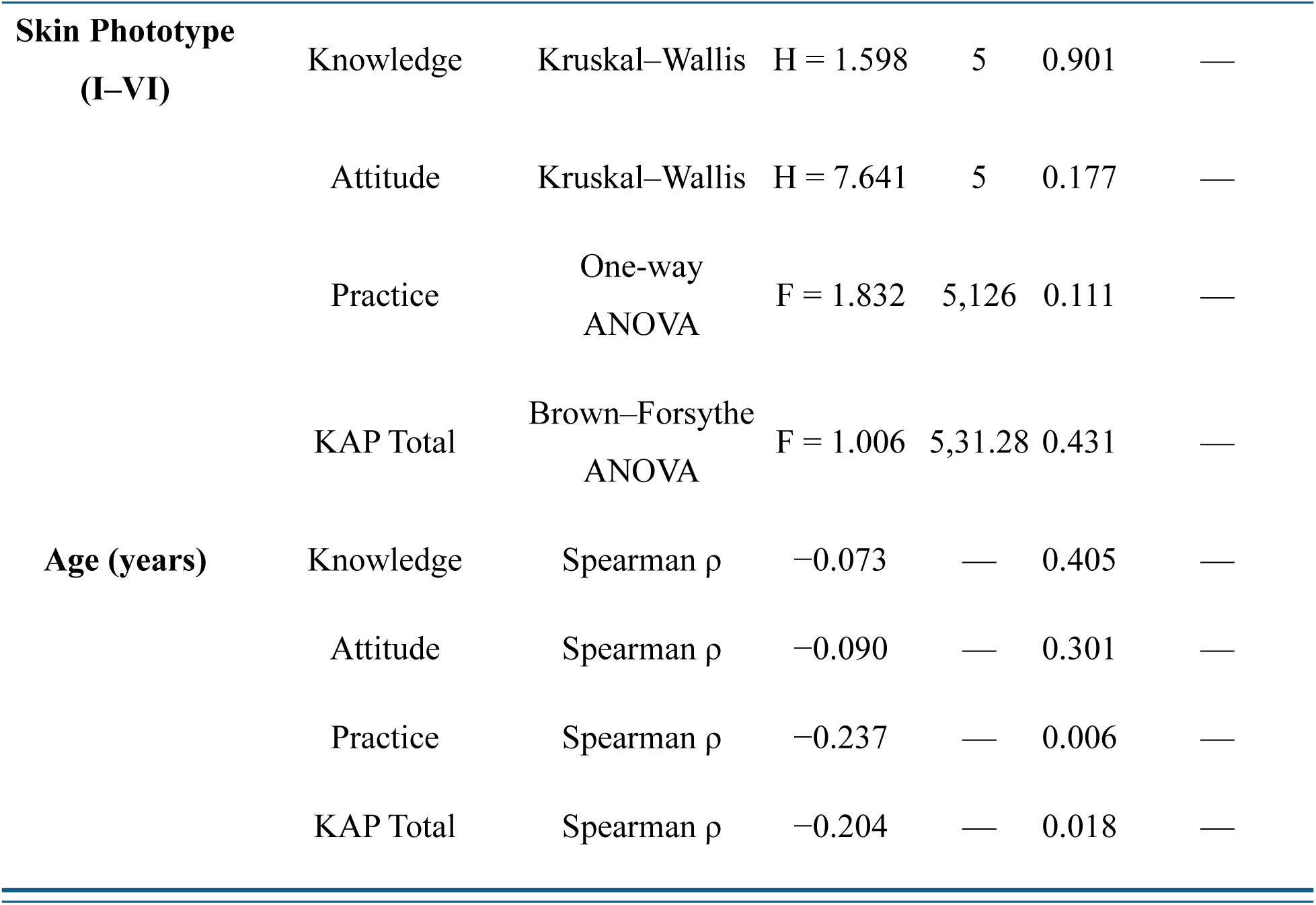
Associations Between Sociodemographics, Photoprotection Practices, and KAP Scores. Data are presented as results of inferential tests. Mann–Whitney U and Kruskal–Wallis tests were applied when normality assumptions were not met. Student’s t-test and one-way ANOVA were used when normality and homogeneity of variances were satisfied. Brown–Forsythe robust ANOVA was applied when variance homogeneity was violated. Spearman’s rank correlation (ρ) was used for age. Statistical significance: p < 0.05.

### 3.5. Barriers and Facilitators

The most frequently reported barrier to sunscreen use was forgetfulness (49.6%), followed by lack of knowledge regarding reapplication frequency (27.8%).

The most commonly reported facilitating factor was the use of reminders (33.8%), followed by improved affordability (21.8%) (Figure 2).

**Figure 2.**
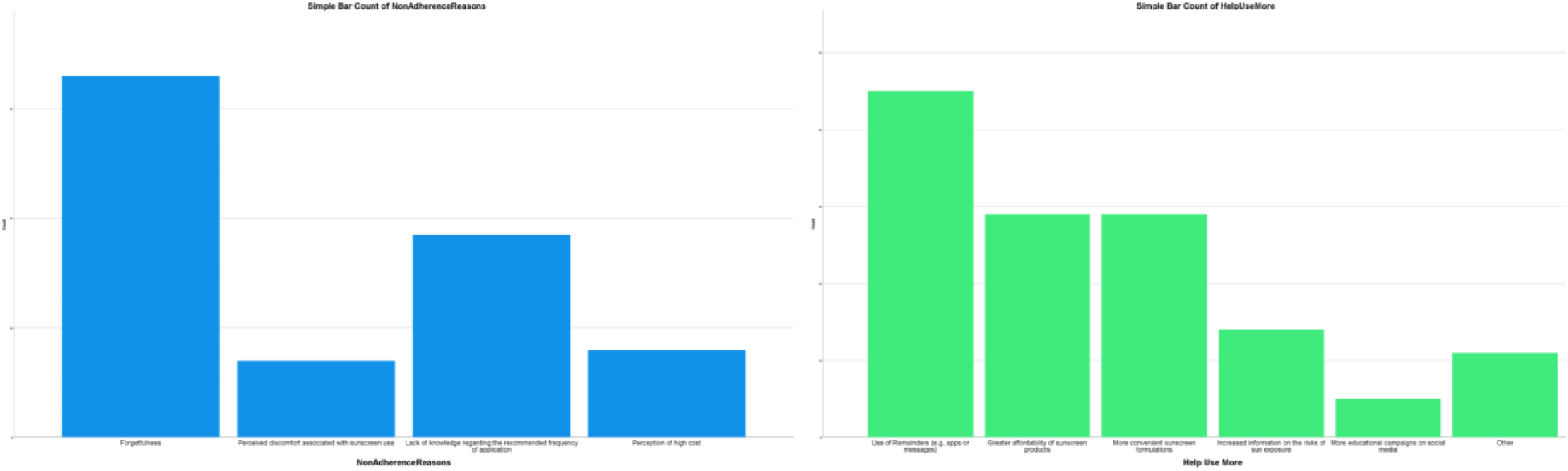
Barriers and facilitators of sunscreen adherence. (Left) Primary reasons for non-adherence to sunscreen use, with forgetfulness being the most prevalent barrier (49.6%), followed by a lack of knowledge regarding reapplication frequency (27.8%). (Right) Factors identified by participants to improve adherence; the implementation of reminders (e.g., apps or messages) was the most frequently cited facilitator (33.8%), followed by improved product affordability (21.8%) and more convenient formulations (21.8%). Data are presented as percentages of the total sample (N = 133).

## Discussion

This study is, to our knowledge, the first to evaluate knowledge, attitudes, and practices (KAP) regarding photoprotection among medical students in Nicaragua and, more broadly, in Central America and the Caribbean. The results reveal a clear mismatch between theoretical knowledge and actual protective behaviors, a pattern consistently described in the international literature [9,13,14].

Overall, students demonstrated acceptable basic awareness. Most reported knowing what sunscreen is (97%) and correctly identified an appropriate product (88%). However, important gaps were evident in key competencies. Only one-third knew the minimum recommended SPF for daily use, and fewer than one-quarter understood endogenous photoprotection mechanisms. These findings indicate that knowledge remains largely superficial. Similar patterns have been reported in Vietnamese students, where high perceived awareness coexisted with low overall knowledge levels [7], and in Mexico, where knowledge scores remained modest throughout medical training, suggesting insufficient curricular emphasis [15].

The discrepancy between knowledge and behavior was pronounced. Although 60.9% reported understanding photoprotection, only 39.1% used sunscreen regularly and 24.8% never used it. Comparable discordance has been observed in multiple settings. In Peru, medical students showed higher knowledge than peers from other disciplines but no behavioral advantage [9]. Spanish data likewise indicate that improved knowledge after dermatology training translated into limited changes in protective habits [13]. Qualitative evidence further shows that students often recognize the risks of sun exposure yet prioritize convenience and comfort over daily sunscreen use [16]. Together, these findings confirm that cognitive awareness alone is insufficient to modify behavior.

Consistent sex differences were observed. Women scored higher across all KAP domains, particularly in practices. This pattern aligns with studies from Turkey reporting more frequent use, better reapplication habits, and greater procedural knowledge among women [14], as well as Brazilian data showing markedly higher non-use among men [17]. However, European evidence suggests a more complex relationship in which women, despite better protective behaviors, may also report greater interest in tanning and longer sun exposure times [8]. Nigerian findings further indicate that gendered marketing of skincare products may contribute to these disparities, highlighting the role of sociocultural and commercial influences [18].

Progression through medical school was not associated with better KAP scores, suggesting that routine training may be insufficient to change behavior. While some studies report incremental improvements in knowledge among senior students, meaningful behavioral change is often limited or delayed [7]. Other research shows that dermatology coursework can improve sunscreen use, although effects vary by context [8]. In contrast, some cohorts demonstrate declining protective behaviors during clinical years, possibly due to competing academic demands [9]. The high frequency of forgetfulness reported in our sample supports this interpretation.

Attitudinal findings revealed both strengths and weaknesses. Most students recommended sunscreen to others, yet only one-third felt adequately informed about sun exposure risks. Objective verification behaviors, such as routinely checking SPF or ingredients, were uncommon. The main barriers identified were forgetfulness and uncertainty about reapplication frequency, consistent with prior reports [9]. Qualitative studies also highlight cosmetic concerns, including greasy texture and difficulty finding suitable formulations, as important but often underestimated determinants of adherence [16]. Encouragingly, students identified practical facilitators such as reminder systems, improved affordability, and more cosmetically acceptable products. Previous intervention studies suggest that even modest educational efforts can improve knowledge and self-examination practices, although sustained change likely requires multifaceted approaches [19].

Important knowledge gaps were also observed regarding environmental impacts of sunscreens and endogenous photoprotection. Similar topic-specific deficiencies have been reported elsewhere [9]. Differences in information sources may partly explain variability across settings. University instruction predominates in some European contexts, whereas dermatologist counseling or internet sources are more influential in others [7,8]. Notably, many students report learning key techniques through social media rather than academic channels, raising concerns about information quality [17].

The phototype distribution reflected Nicaragua’s ethnic diversity, yet no association was found between phototype and protective practices. This suggests that perceived biological risk does not necessarily drive behavior. Evidence from other countries shows wide variation in sunburn prevalence and risk perception [16,17]. In more pigmented populations, the belief that sunscreen is unnecessary remains an important barrier, despite the well-established role of sun exposure in photoaging and pigmentary disorders [13].

The findings have clear implications for medical education and public health. The persistent knowledge–practice gap indicates that information alone is insufficient to change behavior. Effective strategies should promote long-term behavioral autonomy and address sociocultural determinants of sun exposure [14]. Preventive efforts should begin early, particularly given that sunscreen use in this cohort typically started in mid-adolescence [16]. Incorporating reliable educational content into digital and social media environments may further enhance reach and engagement, provided quality is ensured [17].

Improving medical students’ personal photoprotection practices has relevance beyond individual benefit. As future clinicians, they will play a key role in counseling patients on skin cancer prevention and skin self-examination. Strengthening their KAP therefore represents an important investment in the quality of future preventive care [19].

### Limitations

This study has several limitations. Its cross-sectional design prevents establishing causality, and the single-institution sample from an urban area limits generalizability to other medical students in Nicaragua. The use of an ad hoc questionnaire, while necessary for cultural contextualization, means the instrument lacks formal psychometric validation, making comparisons with standardized studies exploratory rather than conclusive [13,15,18]. Additionally, self-reported data are susceptible to social desirability and recall bias, and the sample size fell slightly below the target, reducing statistical power for subgroup analyses. The bivariate analytical approach also cannot account for potential confounding variables.

Although the ad hoc instrument facilitated cultural adaptation, its lack of formal validation restricts direct comparability with studies employing standardized KAP measures. Future research should prioritize the use of psychometrically validated instruments.

Despite these constraints, this study provides valuable baseline data on photoprotection KAP among medical students in a previously unstudied Central American population. By identifying specific knowledge gaps and behavioral barriers, these findings address an important geographic evidence gap and establish a foundation for future research and targeted educational interventions in the region. Finally, cultural norms and values related to sun exposure and skincare in Nicaragua may have influenced participants’ responses and should be considered when interpreting the results.

## Conclusion

This study of photoprotection among medical students in Nicaragua identifies a substantial gap between theoretical knowledge and preventive practice. Although participants demonstrated adequate basic awareness of sunscreen, important deficiencies persisted regarding recommended SPF levels. Notably, fewer than half reported regular sunscreen use despite widespread recognition of its importance, confirming that knowledge alone does not ensure protective behavior. Women achieved significantly higher KAP scores than men, and no improvement was observed across academic years, suggesting that current medical training does not effectively promote sustained behavioral change. The main barriers, particularly forgetfulness and uncertainty about reapplication, are potentially modifiable through focused educational strategies. These findings highlight the need to strengthen practical, behavior-oriented photoprotection training within medical curricula. Future research should incorporate psychometrically validated instruments and longitudinal designs to improve measurement accuracy and enable regional and international comparability. Enhancing medical students’ personal preventive practices is essential for improving future patient counseling and advancing skin cancer prevention in Nicaragua and the broader Central American region.

## Author Contributions

**Jerha Montes:** Conceptualization, methodology, investigation, data curation, formal analysis, writing – original draft, writing – review & editing, final approval of the version to be published, and agreement to be accountable for all aspects of the work.

**Britthany Noguera:** Conceptualization, methodology, investigation, data curation, formal analysis, writing – original draft, writing – review & editing, final approval of the version to be published, and agreement to be accountable for all aspects of the work.

**Ariana Obregón:** Conceptualization, methodology, investigation, data curation, formal analysis, writing – original draft, writing – review & editing, final approval of the version to be published, and agreement to be accountable for all aspects of the work.

**Alejandro Rivas:** Conceptualization, methodology, investigation, data curation, formal analysis, writing – original draft, writing – review & editing, final approval of the version to be published, and agreement to be accountable for all aspects of the work.

**Hallet Whynot:** Conceptualization, methodology, investigation, data curation, formal analysis, writing – original draft, writing – review & editing, final approval of the version to be published, and agreement to be accountable for all aspects of the work.

**Ruby Poveda:** Conceptualization, methodology, validation, investigation, resources, writing – review & editing, supervision, final approval of the version to be published, and agreement to be accountable for all aspects of the work.

**Virgilio Blandón:** Conceptualization, methodology, validation, formal analysis, investigation, resources, writing – original draft, writing – review & editing, supervision, project administration, final approval of the version to be published, and agreement to be accountable for all aspects of the work.

## Acknowledgments

No external contributions, financial support, or material assistance were received for this study.

## Conflicts of Interest

The authors declare no conflicts of interest.

## Data Availability Statement

All supplementary materials supporting the findings of this study, including the complete survey instrument (original Spanish version and English translation), detailed scoring procedures, normality assessments, homogeneity of variance tests, exploratory analyses of individual behaviors, and the systematic literature review with PRISMA flow diagram, are openly available on Zenodo at https://doi.org/10.5281/zenodo.18765924 under the Creative Commons Attribution 4.0 International license (CC BY 4.0). The raw dataset is not publicly available due to privacy concerns, as it contains granular age data that could potentially compromise participant anonymity given the small and localized sample. Researchers seeking access to the data for verification purposes may contact the corresponding author.

The raw dataset containing granular participant data is part of an academic thesis deposited separately in Zenodo and is available under restricted access to protect participant privacy. Researchers seeking access to the dataset for verification or collaborative purposes may request it by contacting the corresponding author. Access will be granted on a case-by-case basis to qualified researchers who provide a reasonable justification. The thesis can be accessed at: https://doi.org/10.5281/zenodo.18530433 (access subject to request/approval).

## Notes

### Competing Interest Statement

The authors have declared no competing interest.

### Funding Statement

This study did not receive any funding

### Author Declarations

Ethics committee/IRB of Universidad Iberoamericana de Ciencias y Tecnologia (UNICIT) gave ethical approval for this work.

